# Policies, practices, opportunities, and challenges for TB screening – A survey of sixty National TB Programmes

**DOI:** 10.1101/2024.07.03.24309601

**Authors:** L Macpherson, C Miller, Y Hamada, MX Rangaka, M Ruhwald, D Falzon, S.V. Kik, H Esmail

**Affiliations:** MRC Clinical Trials Unit at University College London, WC1V 6LJ, United Kingdom; Global TB Programme, World Health Organization, Geneva, Switzerland; WHO Collaborating Centre for Tuberculosis Research and Innovation, Institute for Global Health, University College London, WC1E 6JB, United Kingdom; Centre for Infectious Diseases Research in Africa, Institute of Infectious Disease and Molecular Medicine, University of Cape Town, Observatory 7925, Republic of South Africa; FIND, Geneva, Switzerland

**Keywords:** Tuberculosis, screening, active case finding, subclinical TB, policy

## Abstract

**Background:** To meet incidence reduction goals, the Global Plan to End TB 2023-30 emphasises for the first time that detection of subclinical TB is a priority. WHO Systematic Screening guidelines (2021) have stressed the importance of CXR as a screening tool to achieve this including recommending the use of Computer Aided Detection (CAD) technology.

**Methods:** We conducted a cross-sectional survey of National TB Programmes who reported >1000 TB cases annually. The questions aligned with 2021 WHO screening guidelines and aimed to understand country’s practices, policies, and challenges when screening for TB disease.

**Results:** Sixty of 123 invited countries responded representing 82% of the global TB burden. Only 66% carried out all 6 WHO-recommended steps to implement screening and 39% collected all 7 of the WHO-recommended datapoints for monitoring activity. Although most countries had a policy for using CXR and increasing CXR-based screening (77% and 68% respectively), 90% reported at least one significant barrier to implementing this and 92% reported at least one barrier to implementing CAD technology.

**Conclusion:** Many countries do not carry out all recommended steps for implementation and monitoring of TB screening and although CXR and CAD use are expanding and hold promise as tools to find people with TB, many programmes do not have adequate access to them. While global policy is in place that recommends the use of these tools more efforts should be made to support countries in tackling the barriers that prohibit implementation to make sure that we can close the TB case finding gap.

**What is already known on this topic:** Since the publication of the updated WHO TB screening guidelines there are limited published data on how countries carry out screening for TB disease and what the perceived challenges are for implementing screening from a country perspective.

**What this study adds:** This study provides data on current and planned screening practices and policies within countries as well as the common challenges being faced to implement screening effectively.

**How this study might affect research, practice, or policy:** This information will help developers, policymakers, funding agencies, and academics to better plan and support the roll-out of appropriate screening interventions.

## Introduction

Tuberculosis (TB) continues to be one of the leading causes of death from an infectious disease worldwide, accounting for an estimated 1·3 million deaths in 2022 ^1^. Each year, 4 million cases of TB fail to be diagnosed and/or notified with the potential consequence of uninterrupted community transmission ^1^. This has reaffirmed the focus on systematic screening and early case detection for TB disease as a critical step to ending TB as a global public health challenge within a generation ^1–4^.

Screening for TB disease (as opposed to testing for TB infection) generally relies on an initial evaluation with symptom screen and/or chest X-Ray (CXR) followed by confirmatory testing of those screening positive – usually sputum sampling for bacteriological assessment with molecular assays. However, data from national TB prevalence surveys highlight that most individuals with undiagnosed TB in the community do not report symptoms and thus are missed by typical screening algorithms ^5–8^. Many of these subclinical cases could be detected by CXR ^5,9^. Computer-Aided Detection (CAD) software use deep learning algorithms and artificial intelligence to automate the interpretation of CXRs for signs of TB and several studies have reported equivalence of the diagnostic accuracy of CAD to human readers ^10–12^. These data are reflected in the recently published WHO TB screening guidelines (2021) and Stop TB Partnership’s Global Plan to End TB (2023) ^4,13^. The WHO guidelines highlight the sensitivity of CXR as a screening tool for adults and endorse for the first time the use of CAD software for automated interpretation of digital CXRs when used for TB screening or triage ^13^. The *Global Plan to End TB* emphasises the importance of detecting TB as early as possible, in subclinical stages, by implementing active case finding strategies including use of digital CXR with CAD for interpretation ^4^. However, the potential financial cost and impact of implementing recommended interventions for TB detection are significant for healthcare services. For example, estimates suggest that in 2022 to meet the case detection targets set at the 2018 UN High Level meeting, almost 400 million people would have been needed to be screened for TB disease at a cost of USD 5.3 billion (excluding passive case finding and TB preventive treatment) in high TB burden countries ^14^.

There are limited published data on how countries carry out screening for TB disease and what the perceived challenges are for implementing screening from a country perspective. Understanding these factors are important for developers, policymakers, funding agencies and academics to better plan and support the roll-out of appropriate screening interventions. Following the publication of the updated screening guidelines and subsequent spotlight on TB screening, we conducted a global survey among NTPs. Our aims were to understand countries’ awareness of the updated screening guidelines, to learn more about current and planned screening practices and policies, and to identify the common challenges.

## Methods

### Study design

We conducted a cross-sectional survey with questions that aligned with the 2021 WHO screening guidelines ^2^. There were 26 questions covering three broad themes with relation to TB screening for active disease: 1) *Current policies and practices; 2) Tools and algorithms used; and 3) Barriers and concerns for implementing or expanding X-ray based screening (including the use of CAD)*. The survey was conducted online via Survey Monkey ^15^ with the option to complete a written or verbal version as required. Question styles included multiple choice, checkbox, ranking and free text. The English language survey was translated into Spanish, French, Arabic, and Russian after consultation with WHO head and regional offices. There was no patient or public involvement in the development of this study.

### Data collection and methods

To gather data from countries that represented both a range of stages of the TB epidemic and those who were likely to be carrying out TB screening, we targeted all countries that reported >1000 TB cases per year in 2019. We asked one representative from each NTP to complete the questionnaire, which was developed by the authors with input from the U.K Health Security Agency (UKHSA) and Head and Regional WHO offices. A pilot version was shared with the UKHSA who provided feedback which was incorporated into the final survey to clarify questions. The study methods and results have been reported in keeping with the Checklist for Reporting of Survey Studies (CROSS) ^16^ and a PDF version of the survey can be found in the supplementary materials.

### Survey Administration

Information and the link to the online survey (with an attached PDF) were communicated to one person (in most cases, the lead) from each NTP via email. In the event of no response or a bounced email, a reminder was sent or an alternative contact from within the NTP was sought. The survey was open from December 2021 up to and including March 2022 and during this time a total of 63 individualised email reminders were sent to non-responders. To avoid multiple participation from countries we communicated with one member of each NTP at a time, who could delegate as appropriate. To maximise the number of responders we ensured that the survey took ∼20 minutes to complete, provided a dedicated email address to support participants, engaged with WHO regional offices to support communications, and incentivised countries by hosting a webinar with expert panel to share results and feedback.

### Ethical Considerations

This study had ethical approval from University College London Ethics Committee (19219/001). To maintain anonymity for participants, individual survey responses were not published or shared, instead information was aggregated (either by region or TB burden). The survey responses were also held on a password protected site and password protected computers to protect confidentiality.

### Statistical Analysis

Data was extracted from the online form into an Excel spreadsheet and cleaned. All analysis was performed using Excel (Version 16·75). In the event of missing data i.e., incomplete surveys, respondents were contacted to provide the required information, if this was not successful then missing data was excluded from analysis. We used WHO country groups and World Bank income groups when analysing and reporting results ^17,18^. Respondents were also contacted to clarify any inconsistencies in their responses.

## Results

The survey was sent to 123 countries who reported over 1000 cases of TB in 2019, and 60 (49%) countries responded, representing over 82% of the 2019 global TB incidence (Figure 1) ^19^. Twenty-one (70%) of the 30 high TB burden countries responded to the survey, accounting for 35% of participating countries. The highest response rate was from countries in the Western Pacific WHO region where 67% of the targeted countries responded, followed by 57% from Africa region, 54% from the European region, 36% from South-East Asia, 35% from the Americas, and 29% from Eastern Mediterranean WHO region. Two countries partially completed the survey, and we have stated where these countries have been excluded from analyses. Fifty-three countries responded in English, 5 in French, 2 in Spanish, there were no responses in Arabic or Russian. All countries completed the survey using the online tool. The survey was completed by a senior staff member (director, co-ordinator, manager, or public health specialist) from within the NTP in 93% of cases, the remainder were completed by doctors, officers, or TB advisors from within the NTP.

**Figure 1:**
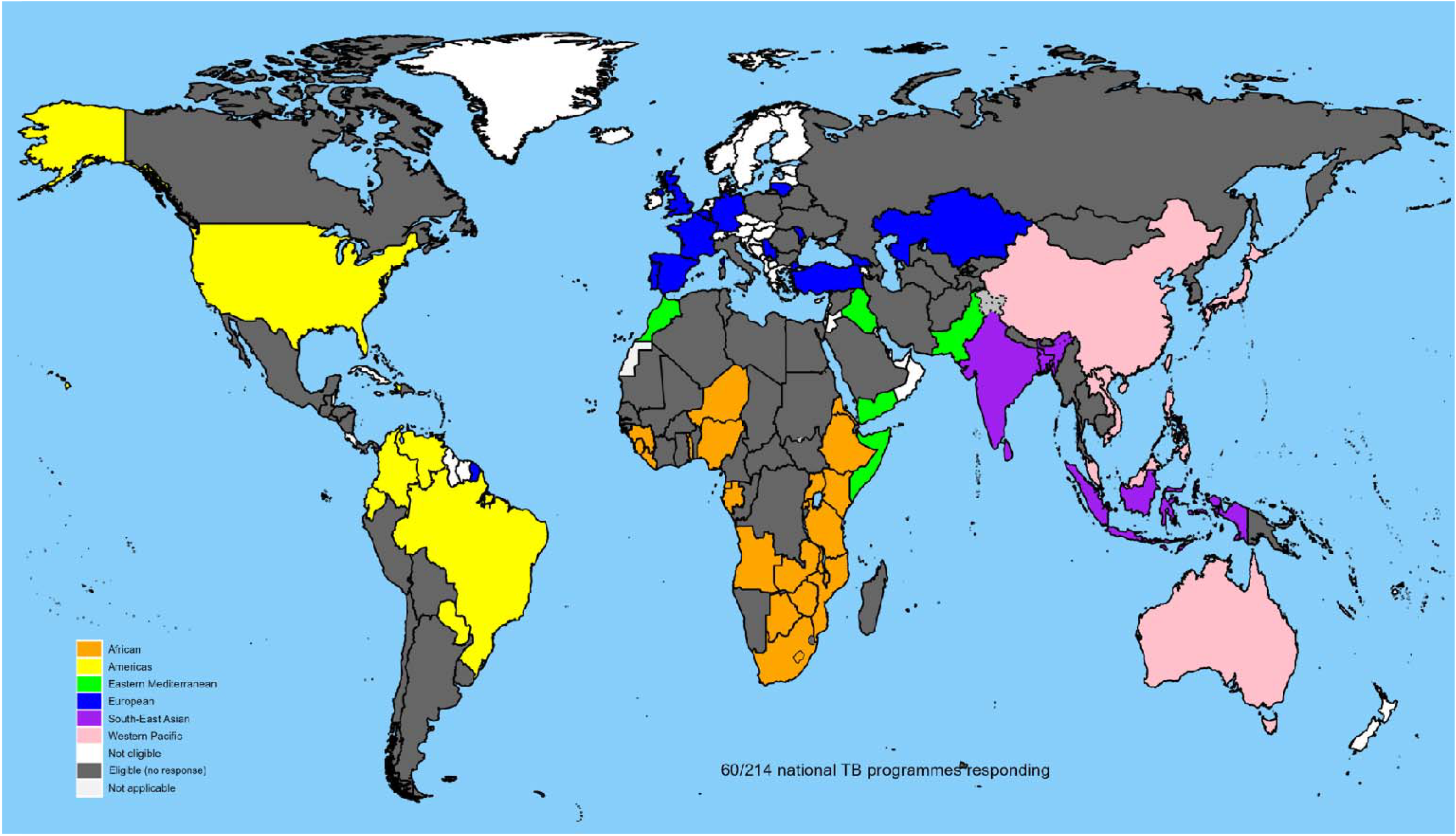
Map showing eligible and participating countries by WHO region. World map showing the participating countries (colour coded by WHO region), eligible countries who reported >1000 TB cases per year in 2019 (coloured grey), and countries who were not eligible to take part in the survey (coloured white).

### Theme 1 – Current policies and practices in relation to screening for TB disease

Fifty-eight (97%) of out of the 60 responders were aware of the updated WHO screening guidelines and 57 (95%) had a national strategic plan for TB. Of these, 50 (88%) planned to increase systematic screening for TB disease, and all but one of the high TB burden countries planned to increase screening. Fifty-seven out of 60 countries (95%) were carrying out screening in one or more high TB risk populations. However, only 66% of countries (39/59 responders), including 55% of high burden countries, carried out all the 6 essential steps outlined by the WHO for designing and implementing a TB screening programme (Figure 2) ^3^.

**Figure 2:**
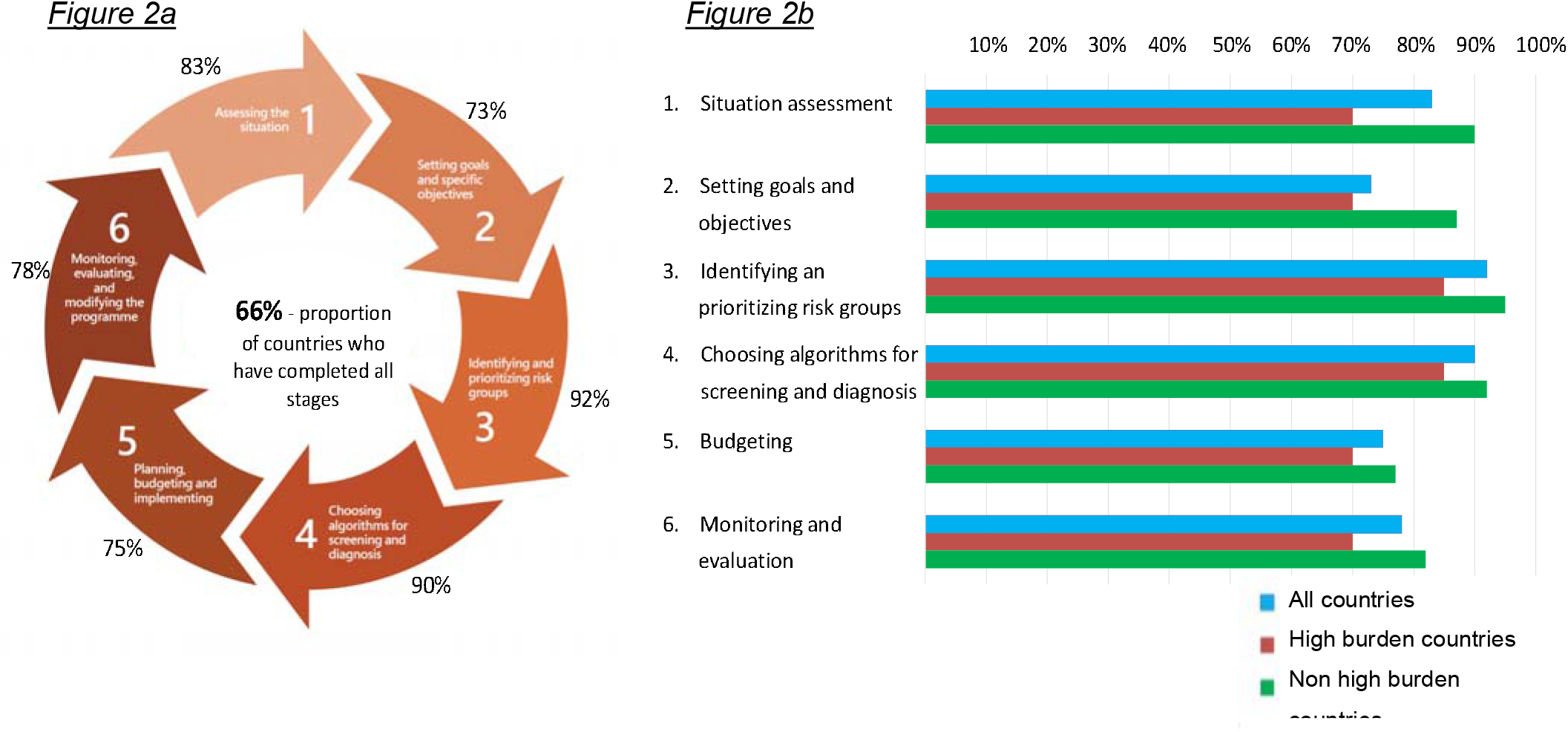
Completion of the six essential steps in the cycle of designing and implementing a TB screening programme (adapted from WHO Operational Handbook on Tuberculosis). This figure contains 2 parts: Figure 2a has been adapted from the WHO Screening Guidelines Operational Handbook and shows the 6 recommended stages of planning and implementing a TB screening programme. The percentages on the outside of the flow chart show the proportion of all countries who reported that they had completed each stage. Figure 2b shows the proportion of countries who had performed each of these 6 stages for all countries and then broken down into countries with high and low TB disease burden.

#### Providers and funding

Three countries (including 2 high burden) reported that they were not performing TB screening, therefore the results in this section are for the 57 remaining responding countries. The public sector provided screening in 98%, and in 71% screening was also being carried out by other bodies including the private sector, non-Governmental organisations (NGOs) or academic institutions. The source of funding for TB screening was domestic in 32% of countries, from international donors in 28% and a combination of the two in 39% (unknown in 2% of countries). Generally, higher-income countries had more domestic funding whereas most (79%) low-income countries relied primarily on international donor funding. For the high TB burden countries, only 2 (11%) reported that screening activities are funded completely domestically.

#### Priority groups

Countries were asked to state whether each of the populations identified by WHO as a risk group for TB was high, neutral, or low priority for TB screening regardless of whether they had a screening policy or were carrying out screening. When prompted, child household contacts, adult household contacts, persons residing in penitentiary institutions, and people living with HIV were all identified by over 80% of countries as high priorities (Figure 3). In addition, over 50% of countries identified people with clinical risk factors for TB (other than HIV) and vulnerable or marginalized groups with limited access to health care as high priority groups. Groups that were identified as low priority for TB screening were communities in remote areas (identified by 23% of countries), urban poor communities (18%), other vulnerable or marginalized groups (17%), internally displaced people (17%), and persons with fibrotic lesions on CXRs (17%). Generally, the high burden countries identified more populations as high priority, with the biggest differences seen for urban poor communities, which 71% of high burden countries identified as high priority compared with 28% from the lower TB burden countries, and communities in remote areas (62% compared with 26%). Migrants were the only population identified as high priority by more lower burden countries - these differences are shown in supplementary Figure 1.

**Figure 3:**
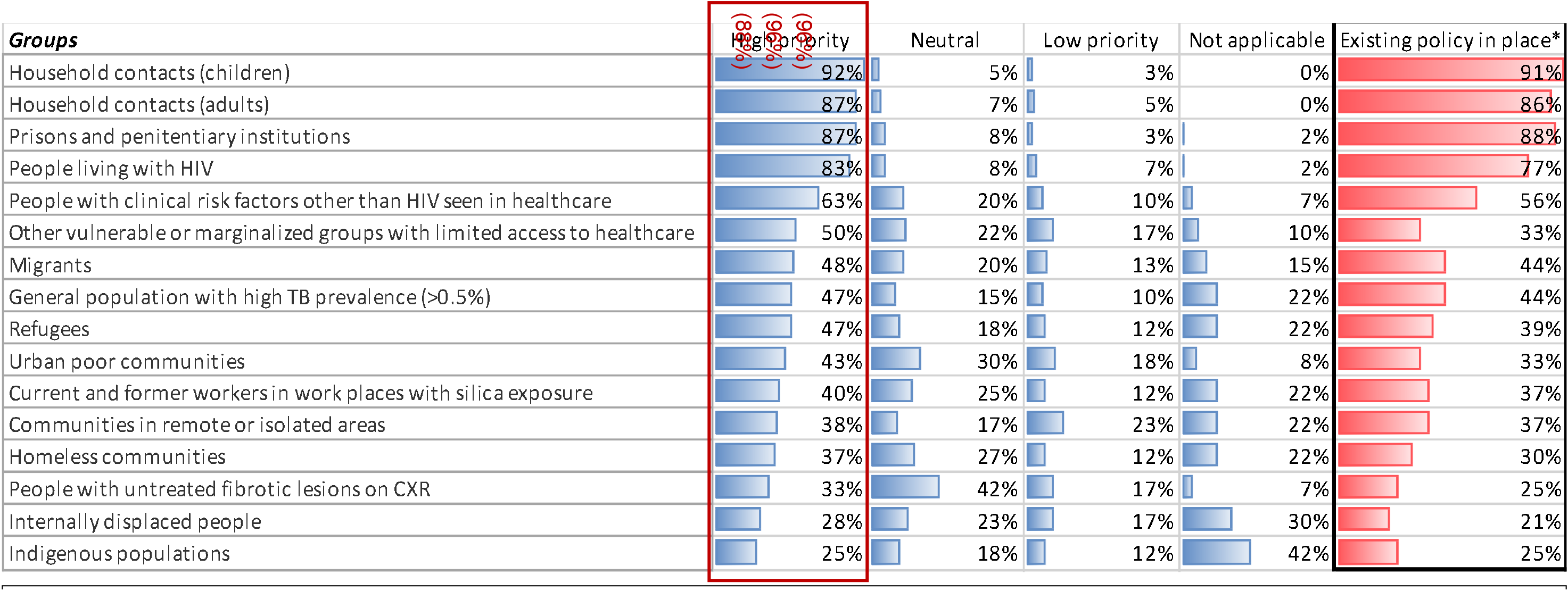
Priority groups for TB screening for all countries. This figure shows the proportion of countries who reported the degree of priority for each risk group identified by WHO. For example, 92% of countries reported that household contacts (children) were of high priority for screening. Countries selected ‘not applicable’ if they felt that the population group was not relevant to their country setting. The column on the right shows the proportion of countries that had an existing policy in place for screening.*Here the denominator is 57 as we excluded those who reported that they were not doing screening. The red box and % show the proportion of countries who had an existing policy for the risk groups that they had identified as high priority. For example, of the 83% who identified people living with HIV as high priority, 88% had a policy to screen in this group.

We also asked whether countries had an existing policy for screening in each of these risk groups. Only 35% of countries (47% of high TB burden countries) had a screening policy for all 4 key risk groups identified by the WHO for whom screening should be performed – household contacts of TB, people living with HIV, persons residing in penitentiary institutions and those with silica exposure. This low percentage is driven by the low proportion of countries (37%) who had a policy for screening those with silica exposure, as most (>77%) did have a policy to screen each of the 3 other key risk groups (Figure 3). For the 4 populations most often identified by participating countries as ‘high priority’ there was an existing policy in place – for example 92% identified persons in penitentiary institutions as high priority and of these 88% had an existing policy for screening in this population. However, there was a lower proportion for whom a screening policy existed for the other population groups, even when identified as high priority (Figure 3).

#### Reporting, monitoring, and evaluation

Forty-two (74%) of the 57 countries who perform TB screening stated that all or most screening activities were reported to the NTP. However, only 22 (39%) countries collected all 7 of the WHO recommended datapoints for monitoring TB screening activity, including 5 of the 19 (26%) high-burden countries. These datapoints span the different stages of the screening process from the number of people eligible for screening to those completing TB treatment. Overall, countries were more frequently collecting the datapoints pertaining to TB diagnosis and treatment (for example 89% of countries report the numbers of people diagnosed with TB) but were less likely to collect data on the numbers of those assessed earlier on in the screening or diagnostic process, for example only 49% of countries reported on the numbers of people eligible for screening (Figure 4).

**Figure 4:**
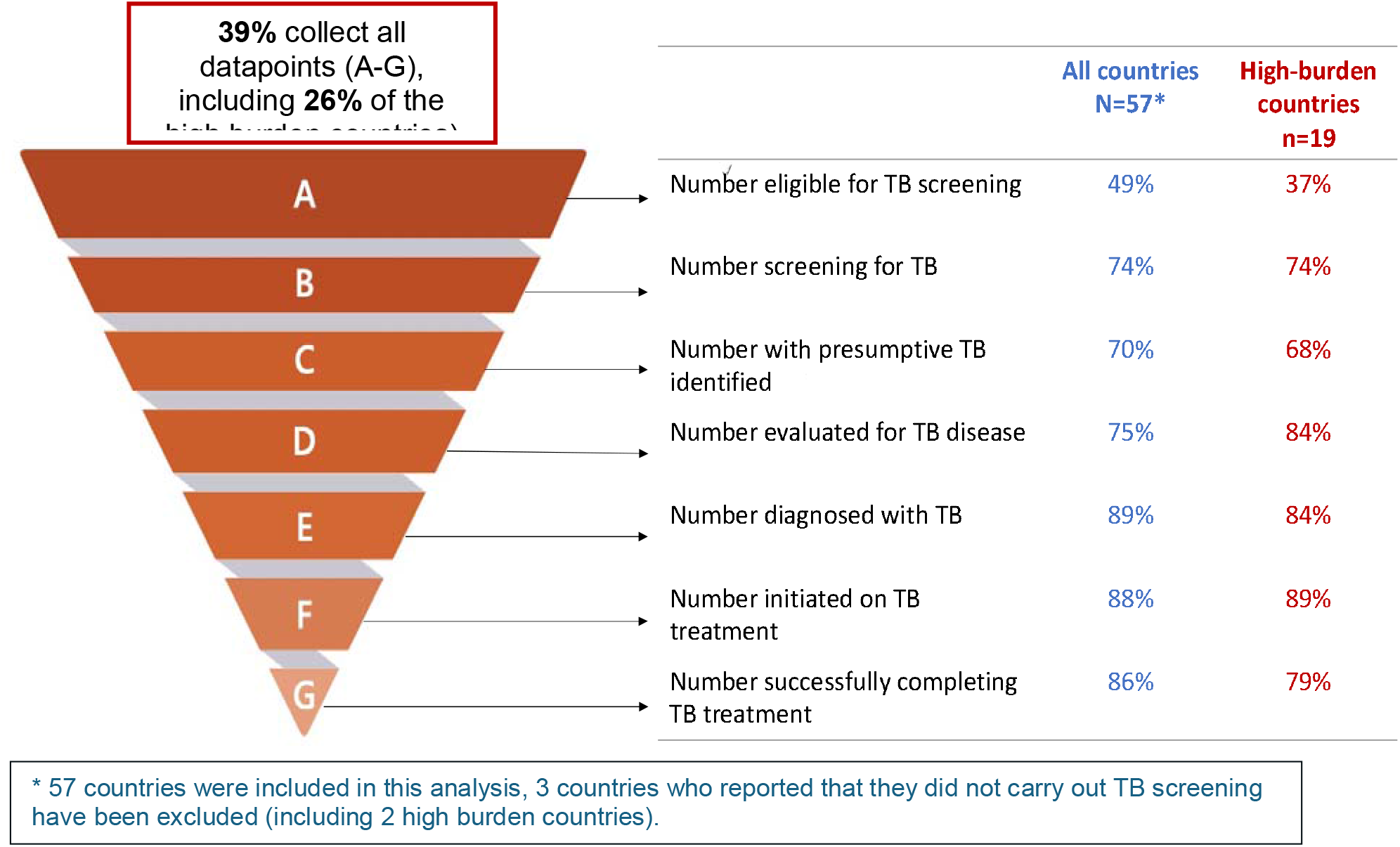
The proportion of countries who collect data for monitoring and evaluation of TB screening as per WHO recommendations. This figure shows the proportion of countries who collect data for monitoring and evaluation of TB screening activity as per the WHO screening guidelines. The inverted pyramid on the left shows the different levels of data that the WHO recommends countries to collect to monitor their screening activity. The table on the right shows the proportion of countries that collect each datapoint, including a separate column highlighting monitoring activity for those high burden countries.

### Theme 2 – Tools and screening algorithms currently used

Participating countries were asked about the screening algorithms they used for 3 risk groups – adult contacts, child contacts and people living with HIV. The data below refer to adult contacts unless otherwise stated and the details for each population group can be found in supplementary Figure 2. Three countries who did not perform systematic screening and 1 country that did not provide answers for this section of the survey were excluded therefore, data for 56 countries including 19 high burden countries are reported below.

### Symptoms and CXR

Forty-six (82%) countries used symptoms to screen for TB disease (cough of any duration, sputum production, haemoptysis, fever, night sweats or weight loss. Of these, 24 (52%) countries also used CXR for *all* those being screened, 11 (24%) only used CXR for those with a positive symptom screen, 3 (7%) only used CXR for those with a negative symptom screen and 8 (17%) did not report any CXR use (supplementary Figure 3). Overall, CXR was used to screen adult contacts for TB by 42/56 (75%) countries including 15/19 (79%) high burden countries and 4/56 countries (7%) reported that they used CXR alone. 5 countries (9%) did not report using either symptoms or CXR (supplementary Figure 3), and 1 country provided inconsistent responses.

Proportionally more high TB burden countries used symptoms to screen (18 (95%)) than non-high burden countries (47 (87%)). Of those using symptoms for screening (n=46) cough of any duration was used more often to screen people living with HIV (used by 87% compared with 48% of countries) (supplementary Figure 2). Fifty-five per cent of countries used the four-symptom-screen (W4SS) to screen people living with HIV. This is the current WHO recommendation for screening in this population (as opposed to using prolonged cough) and comprises screening for current cough, fever, night sweats or weight loss. Overall, 38 (68%) countries including 16 (84%) high TB burden countries plan to implement or expand the use of CXR for systematic screening.

#### CAD

CAD was reportedly used for interpretation of CXRs by 14 (7 high burden) of the 42 countries (33%) who perform CXR-based screening, 4 countries (1 high burden) used CAD for most TB screening, 4 countries (2 high burden) used it in some private sector settings and 6 (4 high burden) used it in some public sector settings. Overall, 16 countries were using CAD in research settings or were planning to trial its use for CXR interpretation, including 8 high burden countries. However, 15 (2 high burden) were not planning to implement CAD over the course of their current national strategic plan.

#### Confirmatory Testing

For confirmatory testing 41/56 countries (73%) used sputum smear, always in combination with culture (31/41 (76%)) and / or molecular (40/41 (98%)) testing. Of the 15 countries who were not using smear, 6 (40%) used culture, 7 (47%) used Xpert MTB/Rif and 9 (60%) used Xpert Ultra (several countries used more than one of these tools). Overall, 37 (66%) countries used sputum culture and 53 (95%) used molecular testing. Of those using molecular testing 41/53 (77%) used Xpert MTB/RIF, 35 (63%) used Xpert Ultra, 3 (5%) used Truenat and 10 (18%) used other molecular tests including line probe assay (often more than one tool was being used).

### Theme 3 – Barriers

Participants were asked about barriers to implementing and expanding CXR use for TB disease screening by selecting from several different options and to provide additional factors. Fifty-four countries provided at least one response to this question, reporting a total of 152 barriers (46% of which were from high burden countries). Of those who responded, most (74%) reported multiple barriers and only 5 (9%) countries reported that there were no barriers (none of which were high burden countries). High equipment costs and/or funding needs were cited most often, selected by 70% of countries and by 90% of the participating high TB burden countries (Figure 5a).

**Figure 5a:**
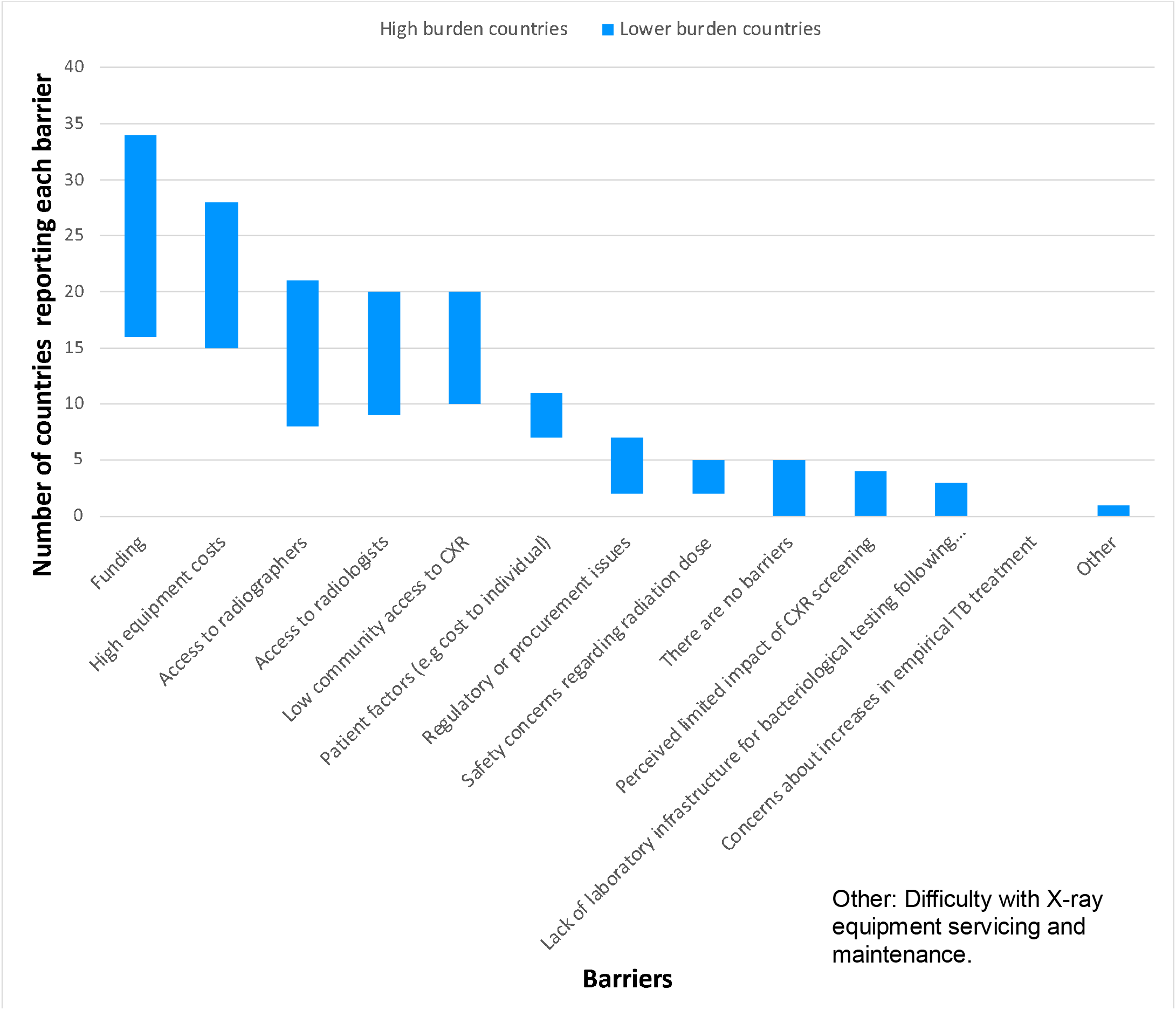
Representation of the barriers to implementing or expanding CXR based screening reported by countries. **Figures 5a and 5b**: These bar charts provide a visual representation of the barriers to implementing CXR based screening (Figure 5a) and CAD for CXR interpretation (Figure 5b) that countries perceive.

With regards to the barriers for implementation of using CAD for the interpretation of CXR, 55 countries provided at least one response, reporting a total of 109 barriers (40% of which were reported by high burden countries). Of those who responded only one (non-high TB burden) country reported that there were no barriers. Eleven countries (20%) reported that they were not aware of the technology, 1 of which is a high TB burden country. The most frequently reported barriers were cost – 18 countries (33%), insufficient infrastructure – 17 countries (31%), connectivity issues – 16 countries (29%), and that there was already sufficient expertise and availability of radiologists for CXR interpretation without CAD – 13 countries (24%) (Figure 5b).

**Figure 5b.**
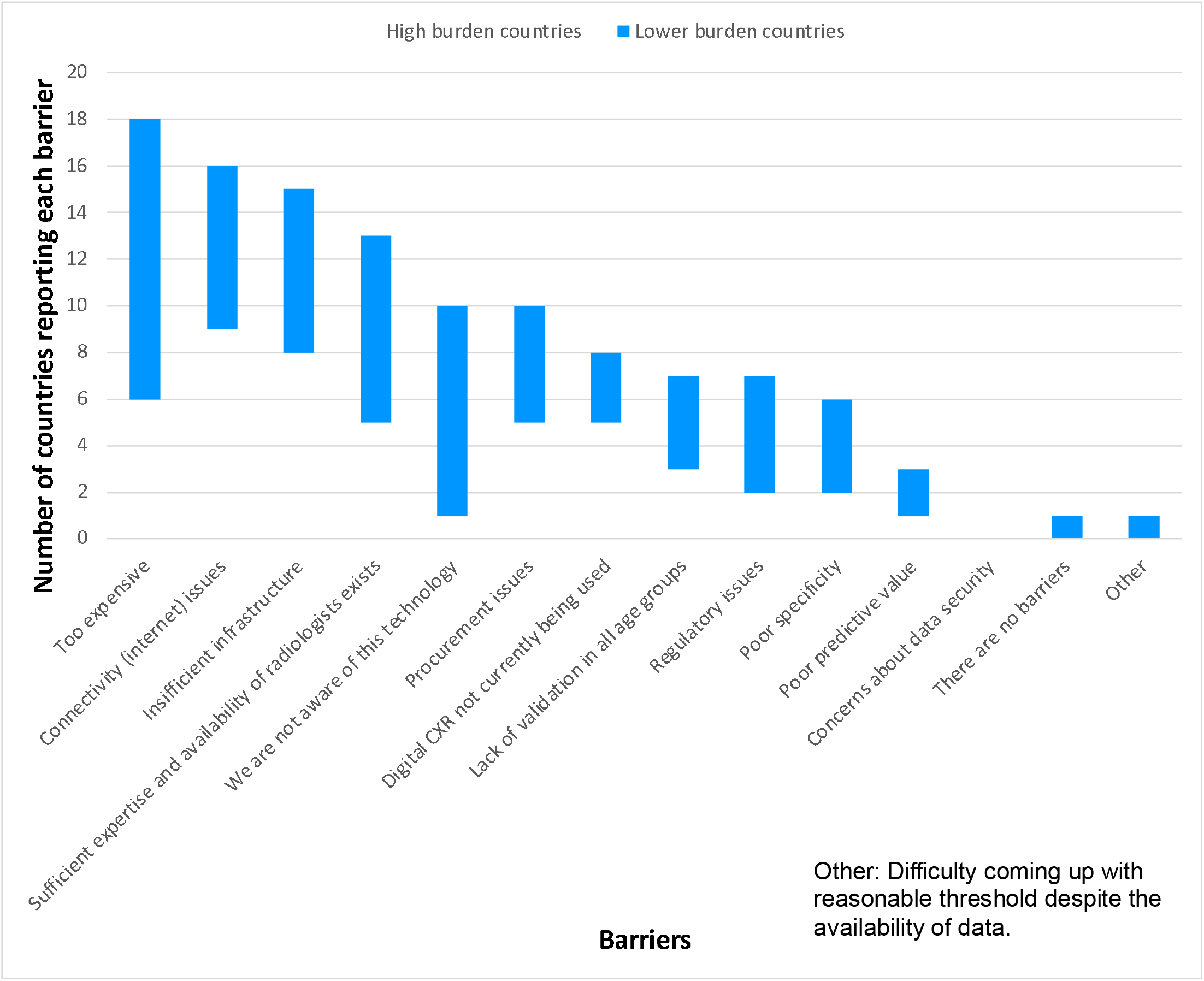
Representation of the barriers regarding implementation of CAD for CXR interpretation in screening for TB disease.

## Discussion

This survey is the largest carried out on screening for TB disease since the publication of the 2021 WHO screening guidelines and the participating countries represents 82% of the global TB incidence. We found that 95% of responding countries had a strategic plan to increase screening for TB disease and that 95% performed TB screening in one or more of the targeted risk groups – most (83%) (particularly the high TB burden countries) used symptom-based screening and many (77%) employed CXR-based screening. Most countries (68%) planned to increase CXR-based screening.

Globally, at a high level, the commitment to screening remains an important aspect of TB elimination – the WHO standard for universal access to rapid tuberculosis diagnostics highlights access to screening and CXR facilities as important benchmarks ^20^. We have shown that this is also true within countries, including many high TB burden countries. However, participating countries reported significant barriers to CXR implementation, in particular funding and resource related. This is consistent with reports from other reviews and surveys which cite factors such as lack of operators, limited access to digital CXR and shortage of radiologists as obstacles ^21–23^.

In our survey over half (55%) of countries who performed CXR-based screening either already used CAD interpretation or planned to pilot its use, although 20% reported that they were not aware of this technology. CAD is likely to become an increasingly important tool for TB screening – several studies report on its equivalent accuracy to human readers for CXR interpretation, it is now endorsed by the WHO screening guidelines, and is also strongly emphasised as a critical tool for case finding in *The Global Plan to End TB* ^4,10–13^. Recently, Barrett and colleagues conducted a survey among TB project implementers who use CXR for TB screening, focusing on CAD interpretation. There were 32 respondents from 19 countries (53% represented NGOs, 19% international organisations, and 13% health facilities), 19/32 respondents used CAD and testified that it enabled them to reach a high throughput, had good accuracy for TB screening, and enormously reduced turnaround times ^22^. There are also organisations working to assist countries with the implementation of CXR and CAD who have created knowledge sharing hubs and make information freely available, allowing implementers to access information about available (portable) digital CXR systems, CAD products and implementation guidance. For example, FIND together with the Stop TB Partnership host the <u>*ai4hlth*</u> resource centre, which features all CAD products, listed by their development and certification stage and can be used for TB detection, the Digital Health Technology Hub from the StopTB Partnership also provides practical guidance on the implementation of CXR and CAD, and WHO TDR provides a research toolkit to support the use of CAD software including calibrating threshold score setting ^24–26^.

### From policy to implementation

Even though 95% of countries were planning to increase TB screening only 66% (and 55% of high burden countries) were carrying out all 6 WHO-recommended steps to implement screening and only 39% (26% of high burden countries) collected all 7 of the WHO-recommended datapoints for monitoring activity. This might reflect the fact that high-burden countries are at capacity with passive case finding (i.e., those presenting to healthcare due to symptoms) and have fewer resources to allocate to active case finding. It also highlights that both implementation and monitoring are areas that would benefit from additional support and investment. This is particularly true in high burden settings who are further away from TB elimination and where health systems are already coping with diagnosing and treating high numbers of TB disease cases and there is less resource to allocate to screening. In these settings the impact of screening is also likely to result in finding more cases compared with lower prevalence settings.

Across the main risk groups, we have shown that >65% of countries used CXR for screening for all participants irrespective of symptoms (supplementary Figure 3) – algorithms that would detect cases of subclinical disease but that are likely to be more expensive due to the cost of increased CXR use as well as greater need for confirmatory testing for those with abnormal (possibly false positive) CXR. A small proportion of countries (7%) used CXR following a negative symptom screen (negative sequential screening) which potentially represents a more cost-effective alternative. Some countries (>15%) used CXR following a positive symptom screen (sequential positive screening) which would be more financially cost effective for the screening programme but would miss those cases who do not report symptoms and might lead to increased costs further down the line. To support the appropriate implementation of these screening tools it will be important to understand more about why countries have chosen specific algorithms and how this will impact the shape of TB prevalence and incidence in their country as well as the economic burden of screening versus missed cases.

### Limitations

This survey was cross-sectional, only answered by one representative from each country, and the results were not checked against any other forms of data or validated using other survey techniques. The questions were mostly multiple choice or easy ‘tick box’ options by design with only a few open-ended questions, making it suboptimal for exploration of context specific details – this was done to ensure that the survey could be completed within an acceptable time frame. Additionally, some questions were open to variable interpretation leading to inconsistencies in our results. For example, in question 14 regarding to what data countries collected for monitoring and evaluation, participants were asked if they collected data on ‘the number of people started on treatment’, this may have been interpreted as overall (i.e. including passive case finding) rather than because of screening alone. We also inferred the types of algorithms countries were using for screening from their answers to the questions on how they used the various tools.

We acknowledge that there may have been significant heterogeneity in TB screening approaches across and within each country and that there likely were intrinsic factors that enabled participation of some countries and not others such as, good internet connectivity, adequate staffing, and willingness to engage. All these factors mean that these data may not be fully representative and limit their generalisability. Despite this, we captured a unique insight into views on TB screening as the countries who did respond accounted for over 80% of the global TB burden.

## Conclusions

Despite 97% of the countries surveyed being aware of the updated screening guidelines many countries – particularly high burden countries – do not carry out all recommended steps for implementation and monitoring of TB screening. Better support in these areas might enable countries to find more cases as meet more of the targets on the way to TB elimination. Only 35% of countries had a screening policy in place for all 4 key risk groups identified by the WHO. CXR and CAD hold promise as screening tool to find people living with subclinical TB when implemented on a larger scale and our data suggest that many countries used CXR in a way that would detect subclinical cases (i.e. for all participants or those with a negative symptom screen). However, according to our survey, many TB programmes do not have adequate access to these tools, due to lack of funding, human resources, or access to information. While global policy is in place that recommends the use of CXR and CAD and where WHO have defined benchmarks for access, more efforts should be made to support countries/NTPs in tackling the barriers that prohibit implementation and large-scale roll-out of these to make sure that we can close the TB case finding gap.

## Supporting information

Survey example

## Data Availability

All data produced in the present study are available upon reasonable request to the authors.

## Acknowledgements

We would like to acknowledge and give special thanks to all those from National TB Programmes around the world who participated in this survey. We would also like to thank our colleagues in the WHO regional offices who helped spread the word and garner enthusiasm for participating in this survey.

The idea for this study was conceived by HE and LM. The survey methods were decided by HE and LM with input from all co-authors. LM collected the data, curated the data, and performed formal analysis with HE. LM prepared the first draft of this manuscript and all co-authors reviewed and edited the manuscript. HE supervised this project.

CM and DF are staff members of the World Health Organization. The authors alone are responsible for the views expressed in this article and they do not necessarily represent the views, decisions or policies of the World Health Organization or other institutions with which they are affiliated.

## Funding

This work was supported by a Medical Research Council award (grant number: MR/V00476X/1) to HE. HE is partially supported by and Medical Research Council unit grant (grant number: MC UU 00004/04).

## Notes

### Competing Interest Statement

The authors have declared no competing interest.

### Author Declarations

This study had ethical approval from University College London Ethics Committee (19219/001).

