## Supplementary materials for "Policies, practices, opportunities, and challenges for TB screening – A survey of sixty National TB Programmes"

Supplementary Table 1: List of eligible countries (reporting >1000 cases per year (2019) invited to participate including those who responded.

Red text indicates WHO high TB-burden country.

Regions: AFR: Africa, AMR: Americas, EMR: Eastern Mediterranean, EUR: European, SEAR: South-East Asia, WPR: Western Pacific.

| Country | WHO Region | Proportion of global TB Burden (%) | Participation |
| --- | --- | --- | --- |
| Algeria | AFR | 0.26 | No |
| Angola | AFR | 1.13 | Yes |
| Benin | AFR | 0.06 | No |
| Botswana | AFR | 0.06 | Yes |
| Burkina Faso | AFR | 0.10 | No |
| Burundi | AFR | 0.12 | No |
| Cameroon | AFR | 0.46 | No |
| Central African Republic | AFR | 0.26 | No |
| Chad | AFR | 0.23 | No |
| Congo | AFR | 0.20 | No |
| Côte d'Ivoire | AFR | 0.35 | No |
| Democratic Republic of the Congo | AFR | 2.80 | No |
| Equatorial Guinea | AFR | 0.03 | Yes |
| Eritrea | AFR | 0.03 | Yes |
| Eswatini | AFR | 0.04 | No |
| Ethiopia | AFR | 1.58 | Yes |
| Gabon | AFR | 0.11 | Yes |
| Gambia | AFR | 0.04 | Yes |
| Ghana | AFR | 0.44 | No |
| Guinea | AFR | 0.22 | Yes |
| Guinea-Bissau | AFR | 0.07 | No |
| Kenya | AFR | 1.41 | Yes |
| Lesotho | AFR | 0.14 | Yes |
| Liberia | AFR | 0.15 | Yes |
| Madagascar | AFR | 0.63 | No |
| Malawi | AFR | 0.27 | Yes |
| Mali | AFR | 0.10 | No |
| Mauritania | AFR | 0.04 | No |
| Mozambique | AFR | 1.11 | Yes |
| Namibia | AFR | 0.12 | No |
| Niger | AFR | 0.20 | Yes |
| Nigeria | AFR | 4.42 | Yes |
| Rwanda | AFR | 0.07 | Yes |
| Senegal | AFR | 0.19 | No |
| Sierra Leone | AFR | 0.23 | Yes |
| South Africa | AFR | 3.62 | Yes |
| South Sudan | AFR | 0.25 | No |
| Togo | AFR | 0.03 | Yes |
| Uganda | AFR | 0.88 | Yes |
| United Republic of Tanzania | AFR | 1.38 | Yes |
| Zambia | AFR | 0.59 | Yes |
| Zimbabwe | AFR | 0.29 | Yes |
| Argentina | AMR | 0.13 | No |
| Bolivia (Plurinational State of) | AMR | 0.12 | No |
| Brazil | AMR | 0.97 | Yes |
| Canada | AMR | 0.02 | No |
| Chile | AMR | 0.03 | No |
| Colombia | AMR | 0.18 | Yes |
| Dominican Republic | AMR | 0.05 | No |
| Ecuador | AMR | 0.08 | Yes |
| El Salvador | AMR | 0.04 | No |
| Guatemala | AMR | 0.05 | No |
| Haiti | AMR | 0.19 | Yes |
| Honduras | AMR | 0.03 | No |
| Mexico | AMR | 0.30 | No |
| Nicaragua | AMR | 0.03 | No |
| Panama | AMR | 0.02 | No |
| Paraguay | AMR | 0.03 | Yes |
| Peru | AMR | 0.39 | No |
| United States of America | AMR | 0.10 | Yes |
| Uruguay | AMR | 0.01 | No |
| Venezuela (Bolivarian Republic of) | AMR | 0.13 | Yes |
| Afghanistan | EMR | 0.72 | No |
| Djibouti | EMR | 0.02 | No |
| Egypt | EMR | 0.12 | No |
| Iran (Islamic Republic of) | EMR | 0.11 | No |
| Iraq | EMR | 0.16 | Yes |
| Libya | EMR | 0.04 | No |
| Morocco | EMR | 0.35 | Yes |
| Pakistan | EMR | 5.73 | Yes |
| Saudi Arabia | EMR | 0.03 | No |
| Somalia | EMR | 0.40 | Yes |
| Sudan | EMR | 0.29 | No |
| Syrian Arab Republic | EMR | 0.03 | No |
| Tunisia | EMR | 0.04 | No |
| Yemen | EMR | 0.14 | Yes |
| Azerbaijan | EUR | 0.06 | No |
| Belarus | EUR | 0.03 | No |
| Belgium | EUR | 0.01 | Yes |
| Bulgaria | EUR | 0.01 | No |
| France | EUR | 0.06 | Yes |
| Georgia | EUR | 0.03 | Yes |
| Germany | EUR | 0.05 | Yes |
| Italy | EUR | 0.04 | No |
| Kazakhstan | EUR | 0.13 | Yes |
| Kyrgyzstan | EUR | 0.07 | No |
| Lithuania | EUR | 0.01 | Yes |
| Poland | EUR | 0.06 | No |
| Portugal | EUR | 0.02 | Yes |
| Republic of Moldova | EUR | 0.03 | Yes |
| Romania | EUR | 0.13 | Yes |
| Russian Federation | EUR | 0.73 | No |
| Serbia | EUR | 0.01 | Yes |
| Spain | EUR | 0.04 | Yes |
| Tajikistan | EUR | 0.08 | No |
| Turkey | EUR | 0.13 | Yes |
| Turkmenistan | EUR | 0.03 | No |
| Ukraine | EUR | 0.34 | No |
| United Kingdom | EUR | 0.05 | Yes |
| Uzbekistan | EUR | 0.22 | No |
| Bangladesh | SEAR | 3.63 | Yes |
| Bhutan | SEAR | 0.01 | No |
| China, Hong Kong SAR | SEAR | 0.05 | No |
| Democratic People's Republic of Korea | SEAR | 1.33 | No |
| India | SEAR | 26.55 | Yes |
| Indonesia | SEAR | 8.50 | Yes |
| Myanmar | SEAR | 1.75 | No |
| Nepal | SEAR | 0.68 | No |
| Sri Lanka | SEAR | 0.14 | Yes |
| Thailand | SEAR | 1.06 | No |
| Timor-Leste | SEAR | 0.06 | No |
| Australia | WPR | 0.02 | Yes |
| Cambodia | WPR | 0.47 | No |
| China | WPR | 8.38 | Yes |
| Japan | WPR | 0.17 | Yes |
| Lao People's Democratic Republic | WPR | 0.11 | Yes |
| Malaysia | WPR | 0.29 | Yes |
| Mongolia | WPR | 0.14 | No |
| Papua New Guinea | WPR | 0.38 | No |
| Philippines | WPR | 6.02 | Yes |
| Republic of Korea | WPR | 0.30 | No |
| Singapore | WPR | 0.02 | Yes |
| Viet Nam | WPR | 1.71 | Yes |

### Supplementary Figure 1: Priority groups for screening for Tuberculosis (TB) disease for high and low TB burden countries.

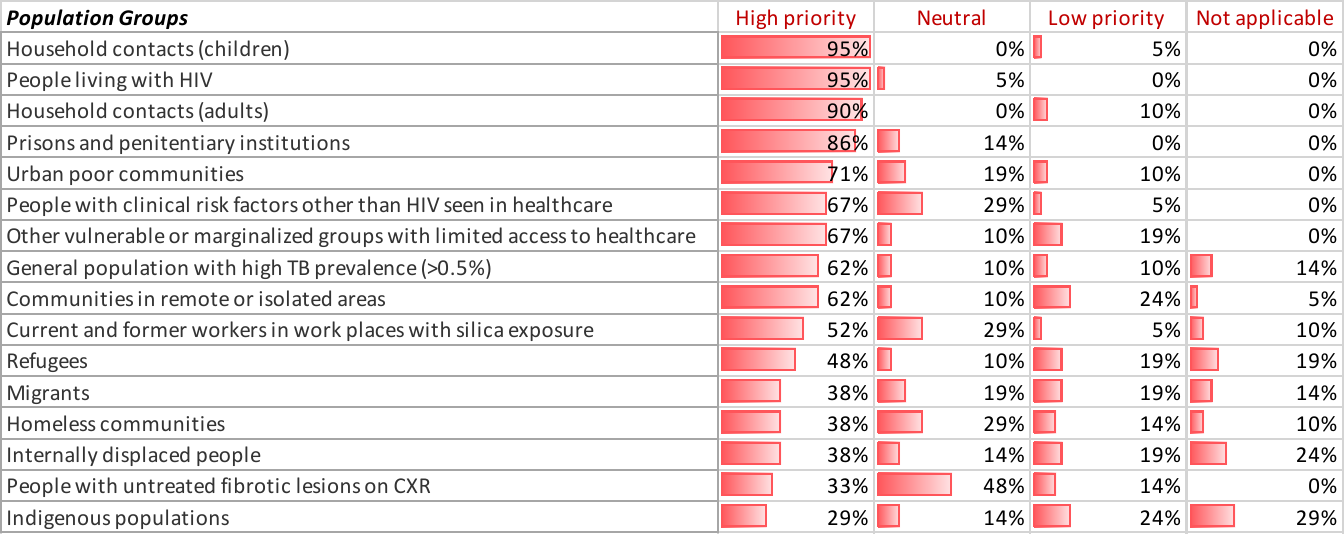
Priority Groups for Screening for Active TB: High TB burden countries (n=21).

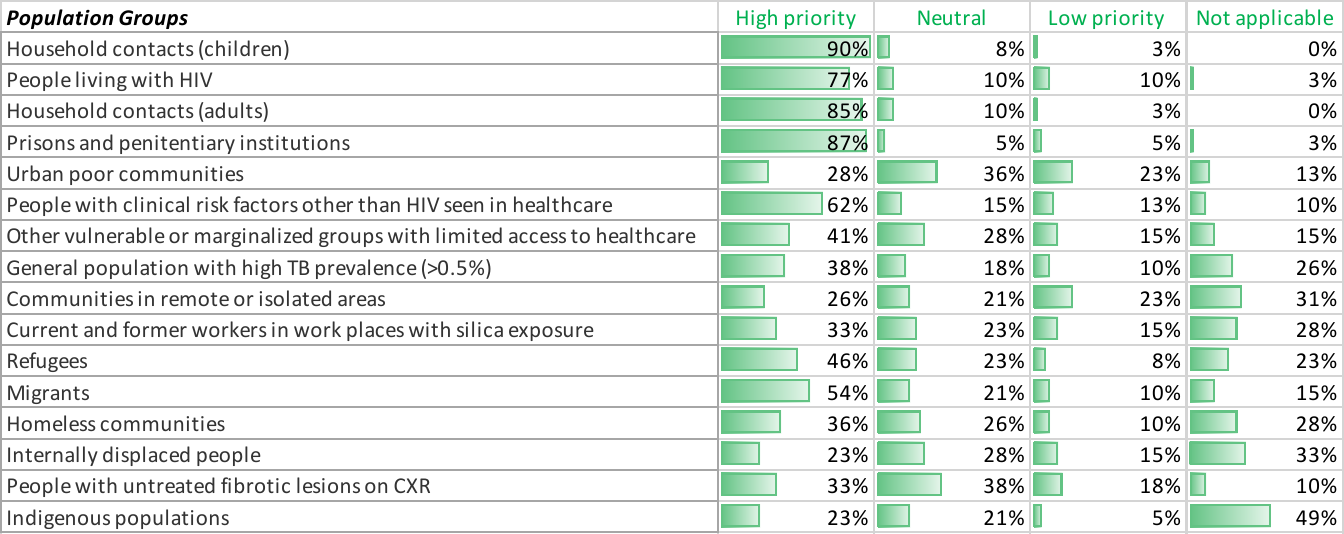
Priority Groups for Screening for Active TB: Lower TB burden countries (n=39).

### Supplementary Figure 2: Representation of the tools reportedly used for Tuberculosis disease screening.

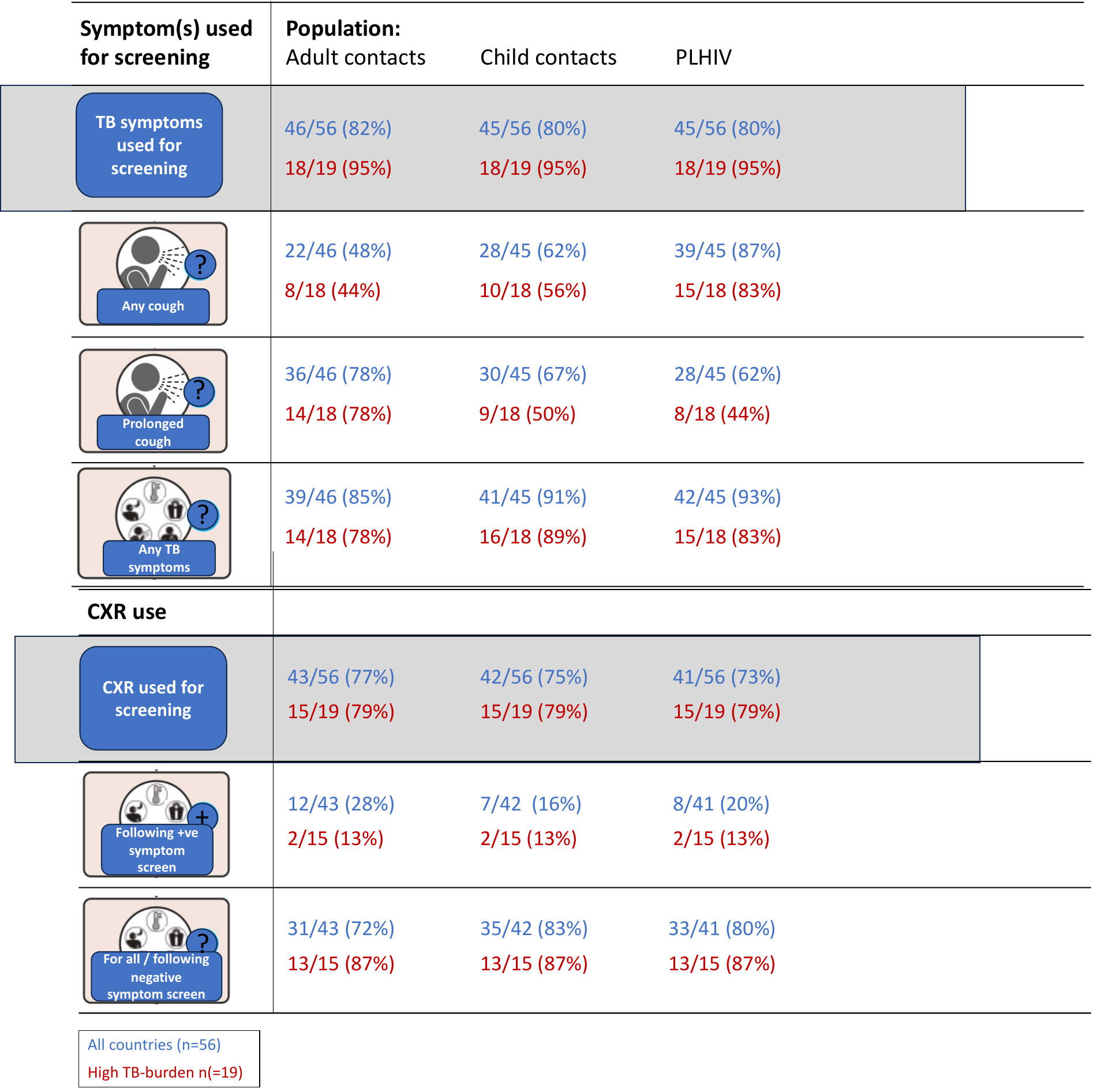

* 4 countries have been excluded: 3 who did not perform systematic screening and 1 that did not provide answers for this section therefore, data for 56 countries including 19 high burden countries are reported here.

*

### Supplementary Figure 3: Screening algorithms used by countries to screen for Tuberculosis disease.

|  | **Number of countries using screening tools (total n=50*)** | | | | |
| --- | --- | --- | --- | --- | --- |
| **Adult contacts** | **8** | **11** | **3** | **24** | **4** |
| **Child contacts** | **9** | **7** | **1** | **29** | **3** |
| **PLHIV** | **9** | **6** | **3** | **28** | **4** |

*Tools used in parallel.*

**Symptom screen**

**CXR**

**Confirmatory testing**

Supplementary figure 4 shoes how countries reported their use of different screening tools for three of the key risk groups – adult TB contacts, child TB contacts and people living with HIV (PLHIV). The yellow box highlights 3 algorithms that would detect subclinical Tuberculosis.

* Exclusions from this figure: 3 countries reported that they did not perform TB screening, 1 did not complete this section of the survey, 1 country provided discrepant answers to these questions, and 5 reported no use of either symptoms or CXR for TB screening.

### Legend.

**Supplementary Table 1: List of eligible countries invited to participate including those who responded.**

Showing a list of eligible countries (who reported >1000 Tuberculosis cases in 2019) who were invited to participate in the survey. The column on the left indicated whether the country participated in the survey.

**Supplementary Figure 1: Priority groups for screening for Tuberculosis disease.**

Showing the priority groups for screening for Tuberculosis disease for both high and lower burden countries.

**Supplementary Figure 2: Representation of tools used for Tuberculosis disease screening.**

This is a visual representation of the types of screening tools used by participating countries to screen for Tuberculosis disease.

**Supplementary Figure 3: Screening algorithms used by countries to screen for Tuberculosis disease.**

This is a visual representation of screening algorithms used by countries to screen for Tuberculosis disease.
