## Supplementary material for "Policies, practices, opportunities, and challenges for TB screening – A survey of sixty National TB Programmes": Survey example

THIS PDF IS FOR REFERENCE ONLY. Please complete the survey online.

#### Introduction

**Thank you for taking part in this global survey on behalf of your country's National TB Programme. The aim is to understand the policies, practices, and challenges of systematic screening for active TB disease in countries that report >1000 TB cases per year. This will provide important information for policymakers, funding agencies and academics involved in maximising the effectiveness of screening for active TB disease.**

**The survey has been developed by a team from University College London, FIND and WHO and has received ethical approval from the University College London Research Ethics Committee. The 2021 WHO guideline on systematic screening for TB disease serves as the benchmark for many of the questions.**

**The questions focus on systematic screening for active TB disease in your country (as opposed to screening for latent TB infection (LTBI)). We estimate that it will take you approximately 20 minutes to complete.**

<u>For the purposes of this survey systematic screening refers to:</u> **The systematic identification of active TB disease in people who are not seeking healthcare, through assessing symptoms and using tests, examinations or other procedures that can be applied rapidly. For those who screen positive, the diagnosis of TB disease then needs to be established by one or several diagnostic tests and additional clinical assessments. This is also known as active case finding and is distinguished from testing for latent TB infection.**

**For any queries please contact:**

THIS PDF IS FOR REFERENCE ONLY. Please complete the survey online.

### Policies, practices, and challenges of systematic screening for TB disease: a global survey.

#### General information

1. Which country's National TB Programme (NTP) are you answering on behalf of?

2. Your job title and affiliation(s)

3. Please state your name and contact details. *This information will only be used to contact you about your survey responses and will not be shared with any third parties.*

**Name**

**Email Address**

**Phone Number**

THIS PDF IS FOR REFERENCE ONLY. Please complete the survey online.

Policies, practices, and challenges of systematic screening for TB disease: a global survey.

#### Policies and practices in relation to systematic screening for TB disease in your country

**The questions in this section focus on your country's policies and practices in relation to systematic screening for active TB disease (not LTBI).**

4. Prior to this survey, were you aware of the 2021 WHO Guidelines on systematic screening for active TB disease?

- ○ Yes
- ○ No

5. Is systematic screening for active TB disease conducted in any form in your country?

- ○ Yes
- ○ No

6. Do you have a national strategic plan for TB

- ○ Yes
- ○ No

If answered "Yes" please state what time period the strategic plan covers

[ ]

7. Does your national strategic plan for TB include a plan to either increase or decrease systematic screening for active TB disease?

- ○ Not applicable (no national strategic plan)
- ○ Increase systematic screening
- ○ Decrease systematic screening
- ○ No plan to increase or decrease
- ○ Don't know
- ○ Other (please specify)

[ ]

8. Do you have an existing national guideline that covers systematic screening for active TB disease?

- ◯ Yes
- ◯ No
- ◯ No - but we have a national policy or guideline in progress
- ◯ Other (please specify)

9. In relation to implementing systematic screening for active TB disease which of the following steps have been carried out or are currently being carried out? *Please answer regardless of whether you have an existing policy for screening. - see chapter 2 of the WHO operational handbook for more information.* [Tick all that apply]

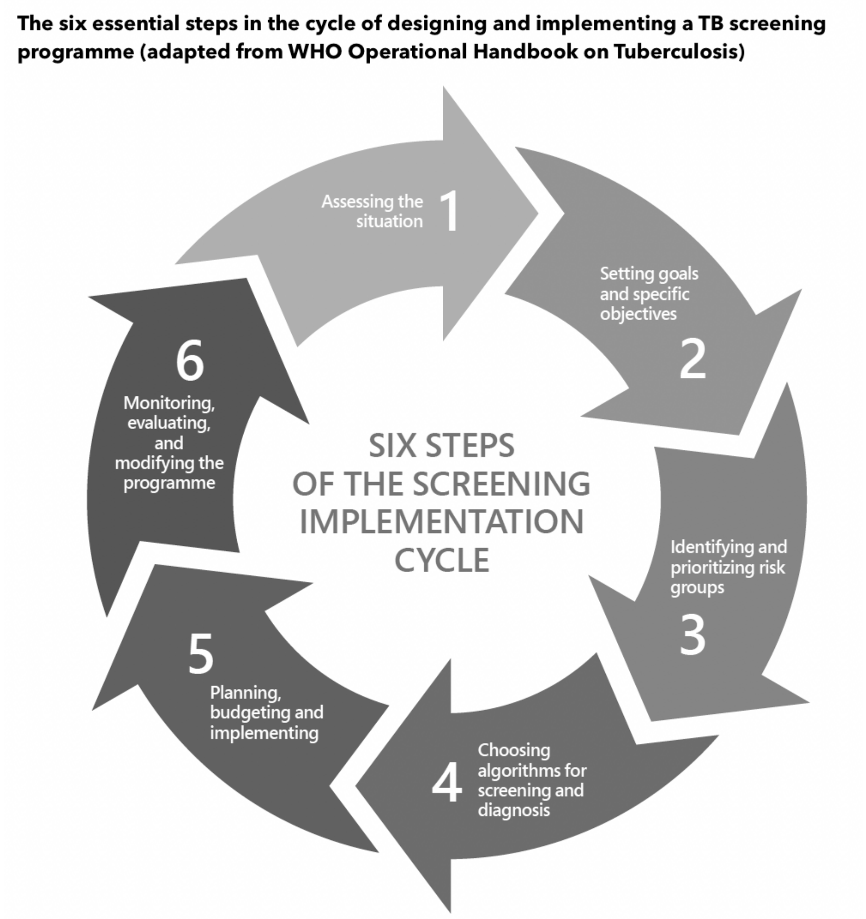


- ☐ Situation assessment
- ☐ Setting goals and specific objectives
- ☐ Identifying and prioritizing risk groups
- ☐ Choosing algorithms for screening and diagnosis
- ☐ Budgeting
- ☐ Monitoring and evaluation
- ☐ None of the above
- ☐ Not applicable

10. Please rate the priority of the following groups for systematic screening for active TB disease in your country? *Please answer regardless of whether you have an existing policy for screening.*

| | Low priority | Neutral | High priority | Not applicable |
|---|---|---|---|---|
| General population with high TB prevalence (>0.5%) | ◯ | ◯ | ◯ | ◯ |
| People living with HIV | ◯ | ◯ | ◯ | ◯ |
| Household contacts (adults) | ◯ | ◯ | ◯ | ◯ |
| Household contacts (children) | ◯ | ◯ | ◯ | ◯ |
| Prisons and penitentiary institutions | ◯ | ◯ | ◯ | ◯ |
| Current and former workers in work places with silica exposure | ◯ | ◯ | ◯ | ◯ |
| People with clinical risk factors other than HIV seen in healthcare | ◯ | ◯ | ◯ | ◯ |
| People with untreated fibrotic lesions on CXR | ◯ | ◯ | ◯ | ◯ |
| Urban poor communities | ◯ | ◯ | ◯ | ◯ |
| Homeless communities | ◯ | ◯ | ◯ | ◯ |
| Communities in remote or isolated areas | ◯ | ◯ | ◯ | ◯ |
| Indigenous populations | ◯ | ◯ | ◯ | ◯ |
| Migrants | ◯ | ◯ | ◯ | ◯ |
| Refugees | ◯ | ◯ | ◯ | ◯ |
| Internally displaced people | ◯ | ◯ | ◯ | ◯ |
| Other vulnerable or marginalized groups with limited access to healthcare | ◯ | ◯ | ◯ | ◯ |

11. In which of the following groups do you currently have a policy for systematic screening for active TB disease? [Tick all that apply]

- [ ] Not applicable (no policy for systematic screening in place)
- [ ] General population with high TB prevalence (e.g. >0.5%)
- [ ] People living with HIV
- [ ] Household contacts (adults)
- [ ] Household contacts (children)
- [ ] Prisons and penitentiary institutions
- [ ] Current and former workers in work places with silica exposure
- [ ] People with clinical risk factors other than HIV seen in healthcare
- [ ] People with untreated fibrotic lesions on CXR
- [ ] Other (please specify)
- [ ] Urban poor communities
- [ ] Homeless communities
- [ ] Communities in remote or isolated areas
- [ ] Indigenous populations
- [ ] Migrants
- [ ] Refugees
- [ ] Internally displaced people
- [ ] Other vulnerable or marginalized groups with limited access to healthcare

12. Which stakeholders or organisations in your country are the main providers of systematic screening for active TB? [Tick all that apply]

- [ ] Government/ public health
- [ ] Private sector
- [ ] Non-Governmental Organisation
- [ ] Academic institution
- [ ] Other (please specify)

13. Approximately what proportion of screening activity for active TB disease, regardless of who provides it, is reported to the National TB programme?

- ◯ Not applicable (no screening activity)
- ◯ All (100%)
- ◯ Majority (75%)
- ◯ Half (50%)
- ◯ Minority (25%)
- ◯ None (0%)

If all screening activity is not reported to NTP please comment on what the challenges are

[ ]

14. What data does your NTP routinely collect for monitoring and evaluating your systematic screening programme for active TB disease? [Tick all that apply]

- ☐ No data is collected for monitoring of systematic screening
- ☐ Number of people eligible for screening according to national guidance
- ☐ Number of people screened for TB (total)
- ☐ Number of people screened by risk group (e.g. contacts, PLHIV, migrants etc)
- ☐ Number of people screened by the screening tool used (e.g. CXR vs symptoms only)
- ☐ Number of people who screen positive for TB (i.e. requiring confirmatory testing)
- ☐ Number of people evaluated for TB disease (i.e. receiving confirmatory testing)
- ☐ Number of people diagnosed with TB (total, including clinical and bacteriological diagnosis)
- ☐ Number of people diagnosed with TB by bacteriological status (i.e. confirmed or not)
- ☐ Number of people diagnosed with TB classified by symptom status
- ☐ Number of people started on TB treatment
- ☐ Number of people successfully completing TB treatment
- ☐ Other (please specify)

[ ]

15. Is systematic screening for active TB disease budgeted in your National TB budget?

- Not applicable (no screening)
- Yes
- No
- Partially
- Don't know
- Other (please specify)

16. What is the source of funding within the National TB budget for systematic screening activity for active TB?

- Not applicable
- Domestic funding
- International donor funding (including the Global Fund)
- Mixture of domestic and international funding
- Unfunded
- Other (please specify)

Please specify other sources of funding and describe the approximate proportion of funding if mixed, for example: 50% domestic funding, 50% international.

17. Going forward, has the priority for systematic screening for active TB changed due to the COVID pandemic?

- Increased priority
- Not affected priority
- Reduced priority
- Don't know
- Other

Please explain your answer

#### Tools and algorithms used for systematic screening for active TB disease in your country

**The questions in this section focus on your country's use of screening tools for active TB disease, focusing on the following population groups: Adult contacts; child contacts; people living with HIV (PLHIV); and other relevant risk groups you feel are screened differently for active TB in your country.**

**For the purposes of this survey the following definitions apply:**
**Prolonged cough: Cough lasting ≥ 2 weeks (please use the comments box to detail any alternative definitions e.g. if your country defines prolonged cough as lasting ≥ 3 weeks).**
**Any TB symptoms: Cough of any duration, sputum, haemoptysis, fever, night sweats or weight loss.**
**W4SS: WHO-recommended four-symptom-screen, comprising screening for a current cough, fever, night sweats or weight loss.**

18. Which symptoms do you use to screen for active TB disease? This applies to the following populations: Adult contacts of TB cases, child contacts of TB cases, PLHIV, and any other relevant risk groups. [Please select 'yes' from the dropdown menu where applicable, otherwise leave the box blank]

| | Adult contacts | Child contacts | PLHIV | Other risk groups |
|---|---|---|---|---|
| Prolonged cough | | | | |
| Any cough | | | | |
| Any TB symptom | | | | |
| W4SS (for PLHIV only) | | | | |
| We do not ask about symptoms | | | | |

Comments (if applicable). For example, if you use symptoms to screen in any other population group or scenario, please provide that information here.

19. Where do you use CXR in your screening algorithm for active TB disease? This applies to the following populations: Adult contacts of TB cases, child contacts of TB cases, PLHIV, and other relevant risk groups. [Please select 'yes' from the dropdown menu where applicable, otherwise leave the box blank]

| | Adult contacts | PLHIV | Children contacts | Other risk groups |
|---|---|---|---|---|
| All people being screened | | | | |
| People with a positive symptom screen (see question 17) | | | | |
| People with a negative symptom screen (see question 17) | | | | |
| People with a positive bacteriological test | | | | |
| We do not use CXR as a screening tool in this context | | | | |

Comments (if applicable). For example, if you use CXR to screen in any other population group or scenario, please provide that information here.

20. Do you use molecular diagnostics (Xpert MTB/RIF, Xpert Ultra or Truenat) as **screening tests** for active TB disease (as opposed to as confirmatory tests)? This applies to the following populations: Contacts of TB cases, PLHIV and any 'other' groups. [Please select 'yes' from the dropdown menu where applicable, otherwise leave the box blank]

| | Contacts | PLHIV | Other |
|---|---|---|---|
| All people being screened | | | |
| All medical inpatients where TB prevalence is high (for PLHIV only) | | | |
| We do not use molecular diagnostics as screening tests for active TB | | | |

Comments (if applicable). For example, if you use molecular diagnostics to screen in any other population group or scenario, please provide that information here.

21. Do you use CRP to screen for active TB disease for PLHIV in your country?

- ◯ Yes
- ◯ No
- ◯ Don't know

Comments (if applicable)

22. What **confirmatory bacteriological tests** for active TB disease are routinely used in people screening positive in your country? [Tick all that apply]

- ☐ Not applicable
- ☐ Smear
- ☐ Culture (MGIT)
- ☐ Culture (solid)
- ☐ Molecular (Xpert MTB/RIF)
- ☐ Molecular (Xpert Ultra)
- ☐ Molecular (Truenat)
- ☐ Molecular (other - please specify in the comments box below)
- ☐ Not stated in guidelines
- ☐ Other (please specify in the comments box below)

Comments (if applicable)

#### The use of CXR technologies for screening for active TB disease in your country

**The questions in this section focus on the use of CXR technologies in your country.**

**For the purposes of this survey the following definitions apply:**

**<u>Computer-Aided Detection (CAD):</u> The use of specialised software to interpret abnormalities on digital chest radiographs that are suggestive of TB. The results are expressed as abnormality scores.**

23. Is implementing or expanding the use of CXR for systematic screening for active TB disease part of your national strategic plan?

- ◯ Yes
- ◯ No
- ◯ Don't know

24. What are the barriers to implementing or expanding CXR based systematic screening for active TB disease in your country? *Please answer regardless of whether you have an existing policy for screening.* [Tick all that apply]

- ☐ High equipment costs
- ☐ Funding
- ☐ Access to radiologists
- ☐ Access to radiographers
- ☐ Lack of laboratory infrastructure for bacteriological testing following CXR
- ☐ Regulatory or procurement issues
- ☐ Perceived limited impact of CXR screening
- ☐ Concerns about increases in empirical TB treatment
- ☐ Patient factors (e.g cost to individual)
- ☐ Safety concerns regarding radiation dose
- ☐ Low community access to CXR
- ☐ There are no barriers
- ☐ Other (please specify)

25. If CXR is used in systematic screening for active TB, is CAD used to inform management decisions?

- ◯ Not applicable - not using CXR for screening
- ◯ No - using CXR from screening but not using CAD
- ◯ No – but planning to pilot use of CAD
- ◯ Only using in research settings or as a pilot
- ◯ Yes - using in some private sector settings
- ◯ Yes - using in some public sector settings
- ◯ Yes - in most settings for TB screening

Comment (if applicable)

26. What are the barriers and concerns regarding implementation of CAD in screening for active TB in all settings? [Tick all that apply]

- ☐ We are not aware of this technology
- ☐ Digital CXR not currently being used
- ☐ Lack of validation in all age groups
- ☐ Sufficient expertise and availability of radiologists exists
- ☐ Too expensive
- ☐ Poor specificity
- ☐ Poor predictive value
- ☐ Regulatory issues
- ☐ Connectivity (internet) issues
- ☐ Procurement issues
- ☐ Insufficient infrastructure
- ☐ Concerns about data security
- ☐ There are no barriers
- ☐ Other (please specify)

THIS PDF IS FOR REFERENCE ONLY. Please complete the survey online.

Policies, practices, and challenges of systematic screening for TB disease: a global survey.

Thank you for taking the time to complete this survey.

27. Are you happy for us to contact you about your answers?

- ◯ Yes
- ◯ No
